# Progression of COVID19 Pandemic in India: A Concurrent Linear Regression Analysis Approach

**DOI:** 10.1101/2021.06.01.21258138

**Authors:** Aalok Ranjan Chaurasia, Brijesh P. Singh, Ravendra Singh

## Abstract

This paper uses concurrent linear regression analysis approach to describe the progression of COVID 19 pandemic in India during the period 15 March 2020 through 15 May 2021. The approach provides very good fit to the daily reported new confirmed cases of the disease. The paper suggests that, based on the parameter of the model, an early warning system may be developed and institutionalised to undertaken necessary measures to control the spread of the disease, thereby controlling the pandemic.

## Introduction

The COVID 19 pandemic in India has been subject to intensive modelling of the progression of the pandemic in the country. These studies have followed both epidemiological and statistical approaches. The epidemiological modelling studies are based on the basic Susceptible-Infected-Recovered (SIR) model and its numerous extensions. Statistical approaches, on the other hand, have followed time series analysis using ARIMA and other models. The focus of these studies has generally been on forecasting the progression of the pandemic, especially, when the peak of the pandemic will arrive and when new infections of Novel Coronavirus will start decreasing. These studies have generally not attempted to describe the progression of the pandemic other than that the appropriateness of the model in fitting the past trend. One limitation that may be found in all these studies is that they have followed a piece-meal approach in analysing the progress of the pandemic characterising the pandemic in the country in terms of waves. The progression of the pandemic in India is generally described in two waves, the first beginning March 2020 and the other beginning March 2021. To the best of our knowledge, there is no study that has analysed the progression of the pandemic right since the first outbreak of the pandemic in the early 2020 taking into the account both waves of the pandemic. Recently, many studies have focused on the second wave of the pandemic only without any consideration to the progression of the pandemic prior to the second wave. Such an approach contributes little to the understanding of the progression of the pandemic and the possible factors behind changes in the pandemic curve.

There are many techniques and tools have been used to understand the past and the future progression of the pandemic (Batista, 2020; Koo et al. 2020; Kucharski et al. 2020; Tuite et al. 2020 and Wu et al. 2020). Wu et al (2020) have applied the stochastic Markov Chain Monte Carlo method while Zhao et al (2020) applied the statistical exponential growth model to describe the progression of the pandemic. Shen (2020), on the other hand, a three-parameter logistic growth function to predict the progression of the pandemic in China and in some other countries. In India, Malhotra et al. (2020) applied the SIR and logistic growth models forecast the progression of the pandemic in the country and in its constituent states. Different mathematical models have also been applied to describe the progression of the pandemic in the country (Rao et al, 2020; Chang et al, 2020; Chaurasia and Singh, 2020; Singh, 2020). Other studies (Singh and Singh, 2020; Huang et al, 2020; Hui et al, 2020; Corman et al, 2020; Rothe et al, 2020; Anastassopoulou et al, 2020; Gamero et al, 2020) have modelled the progression of the pandemic based on different principles of mathematics. Chatterjee et al (2020) used a stochastic mathematical model while Sharma and Nigam (2020), Swain et al (2020) and Newtonraj et al (2020) applied ARIMA model to describe the progression of the pandemic. Mishra et al (2020) advocated SARIMA along with ARIMA for modelling and forecasting the pandemic in India but the study was confined to the first phase of the pandemic only. Bedi et al (2021) used SEIRD and LSTM models but this study was also limited to the first wave of the pandemic.

In this paper, we follow a data driven approach to describe the progression of the COVID-19 pandemic in India since March 2020. The underlying assumption of our analysis is that COVID-19 is transmitted from individual to individual only. This means that the reported number of new cases of the disease on a particular day is related to the reported number of new cases of the disease during the immediately preceding period. A person infected with Novel Coronavirus remains infectious, capable of infecting others on contact, for a period of 9-21 days after catching the infection. It is, however, argued that there is little chance that an infected person can infect other persons in contact after a period of 14 days. In other words, the progression of the COVID-19 pandemic may be described in terms of the relationship between the reported numbers of new cases of the disease on a particular day with the number of new cases of the disease reported during the immediately preceding 14 days period. We found on the basis of the daily reported new cases of the disease that out approach has been able to describe the peaks and troughs in the progress of the pandemic in the country right since March 2020. The analysis also explains the factors responsible for the second wave of the pandemic that the country has witnessed recently and that has been quite severe.

The paper is organised as follows. The next section describes the data used. The only source of data about the progress of the pandemic is the daily reported number of new confirmed cases of COVID-19 that is regularly released by the Government of India. These cases are those who have been tested positive for the Novel Coronavirus after testing in the laboratory. The third section of the paper describes the method used for describing the progression of the pandemic. We have used the functional concurrent linear regression analysis approach or the varying coefficients regression approach to describe the progression of the pandemic in the country. The fourth section of the paper presents the key findings of the analysis. The implications of the findings of the analysis from the point of view of the management and control of the pandemic have been discussed in the fifth section of the paper. The last section of the paper summarises the main findings of the analysis and calls on institutionalising an early warning system based on the application of the model.

### Data

The analysis is based on the daily reported number of confirmed cases of the disease released by the Government of India daily and publicly available on the website www.covid19india.org. These cases are those which have been tested positive for the Novel Coronavirus. They do not include symptomatic cases who have not been tested positive for the Novel Coronavirus. An examination of the data released by the Government of India suggests that there are inconsistencies of the unknown origin in the daily reporting of new confirmed cases of the disease. More-specifically, there is a dip in the number of new confirmed cases of the disease on Monday of almost every week. It is argued that this dip in the new confirmed cases of the disease is due to the decrease in the number of persons tested for Novel Coronavirus carried out on Sunday because many testing facilities remain closed on Sunday as well as because of many other reasons such as closure of the market and business activities. In an attempt to minimise these and other errors associated with the reported number of new confirmed cases of the disease, the present analysis is based on 7-days moving average of the reported number of new confirmed cases of the disease released by the Government of India.

## Method

In what follows, let *Y* denotes 7-days moving average of the number of daily reported confirmed cases of COVID-19. Let the entire period of pandemic is divided into concurrent time segments of 14-days each so that *Y*_*jk*_ denotes the number of new confirmed cases of the disease reported on day *j* of the time segment *k, j* =1, 2,…14. We assume that the number of new confirmed cases of the disease reported during time segment *k* +1 may be related to the number of new confirmed cases of the disease reported during the time segment *k* by the following relationship:

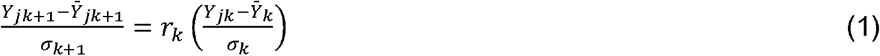

Where *r*_*k*_ is the correlation coefficient between *Y*_*jk*_ and *Y*_*jk* +1_, *j* =1, 2,…14, 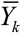 denotes the average of the number of new confirmed cases of the disease reported during the time segment *k* while *σ* _*k*_ is the standard deviation. Equation (1) can be rewritten as,

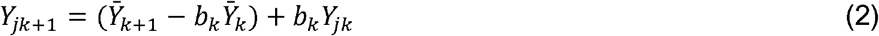

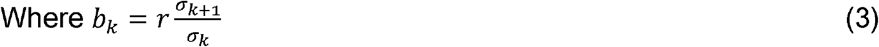

Equation (3) suggests that when *Y*_*jk* +1_ = *b*_*k*_*Y*_*jk*_ for all 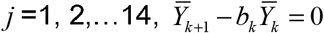. In this ideal situation, number of new confirmed cases of the disease reported during the time segment *j* +1 is *b*_*k*_ times the number of confirmed cases of the disease reported in the time segment *k*. This essentially means that all new confirmed cases of the disease reported in the time segment *k* + 1 are known contacts of the number of new confirmed cases of the disease reported in the time segment *k*. It is, however, rare that 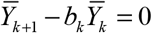 for two reasons. First, all known contacts of the new confirmed cases of disease reported during the time segment *k* who may have been infected because of the contact with an infected person may not be reported during the time segment *k* + 1. Second, some of the new confirmed cases of the disease reported during the time segment *k* + 1 may be of unknown contact. It may be noticed that when 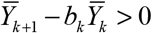, there are some new confirmed cases of the disease reported in the time segment *k* + 1 who are of unknown contacts. On the other hand, when 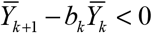, there are some known contacts of the new confirmed cases of the disease reported during the time segment *k* who may have not been reported as new confirmed cases of the disease during the time segment *k* +1 subject to the inconsistencies associated with the daily reporting of new confirmed cases of the disease.

Model (2) suggests that progression of the pandemic can be characterised in terms of the parameter *b*_*k*_. When *b*_*k*_ > 1, *Y*_*jk* +1_ > *Y*_*jk*_, *j* =1, 2,…14 which means an increase in the number of new confirmed cases of the disease during the time segment *k* + 1 relative to the number of new confirmed cases of the disease reported during the time segment *k*. On the other hand, when *b*_*k*_ < 1, *Y*_*jk* +1_ < *Y*_*jk*_, *j* =1, 2,…14, there is a decrease in the number of new confirmed cases of the disease during the time segment *k* +1 relative to the number of new confirmed cases of the disease reported during the time segment *k*. If all contacts of the new confirmed cases of the disease reported during the time segment *k* are traced and tested, then the number of new confirmed cases of the disease reported during the time segment *k* +1 can be determined by the number of new confirmed cases of the disease reported during the time segment *k*.

Model (2) is an example of linear concurrent model in which the response at a given point is modelled as a function of the values of the explanatory variables only at that particular point. It assumes a linear relationship between the reported number of new confirmed cases of the disease with the number of new confirmed cases of the disease reported during the immediately preceding period of days. These models are also termed as varying coefficient models as the parameters of the model change with time so that it follows a trend and is not constant. The linear concurrent models have been found to be useful in the analysis of longitudinal data as is the case here. many settings such as the case here (Brumback and Rice, 1998; Cleveland et al, 1991). Model (2) is characterised by only one parameter *b*_*k*_. It can, however, be extended to include more explanatory variables.

## Results

Figure 1 presents the trend in the parameter *b*_*k*_ of model (2) beginning the time segment beginning 15^th^ March 2020 which gives an idea about the progression of the pandemic. The trend is highly volatile. During its early days, the pandemic in the country progressed very rapidly as is reflected from vary high values of *b*_*k*_. For example, number of new confirmed cases of the disease reported during the 14-days period beginning 17^th^ March 2020 was more than 17 times the number of confirmed cases of the disease reported during the 14-days period beginning 3^rd^ March 2020. This very rapidly progressing first phase of the pandemic appears to have lasted till the time segment beginning 30^th^ March 2020. During this phase, the number of confirmed cases of disease has multiplied very rapidly.

**Figure 1:**
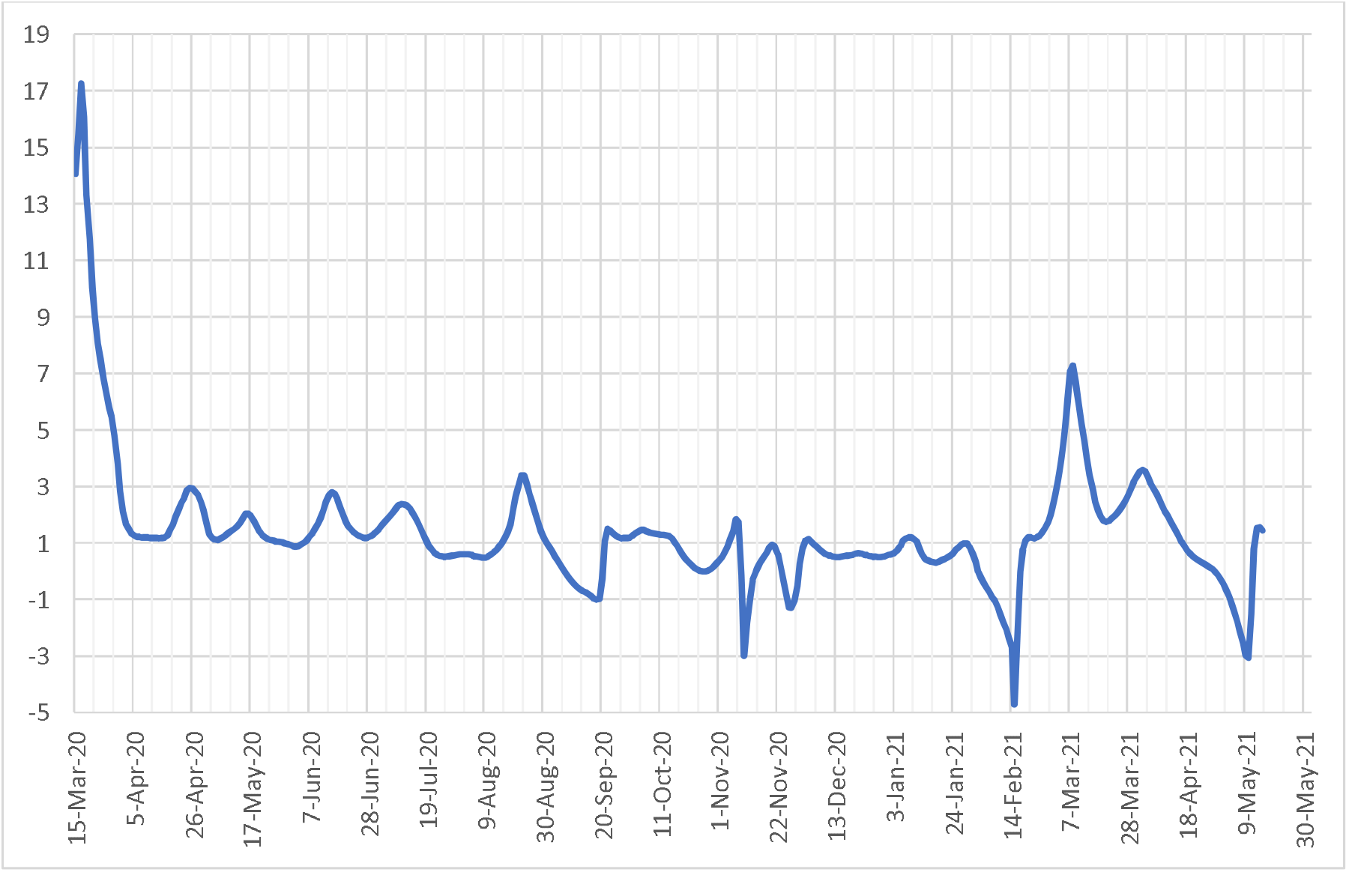
Trend in the parameter b_k_. Source: Authors’ calculations

The second phase of the pandemic appears to have started around the time segment beginning 31^st^ March 2020 and lasted till the time segment beginning 19^th^ July 2020. During this period, the parameter *b*_*k*_ of the model (2) has always been more than 1 but less than 2 except for a few days when it was less than 1 and for a few days when it was more than 2 but always less than 3. This means that, except for a few days during this period, there has been an increase in the reported number of new confirmed cases of the disease during the 14-days period *k*+1 relative to the 14-days period k. However, the pace of the increase in the reported number of new confirmed cases of the disease was not fast as the parameter *b*_*k*_ was, in general varied between 1 and 2 and was rarely more than 2.

The third phase of the pandemic appears to have started around the time segment beginning 20^th^ July 2020 and lasted up to the time segment beginning 15^th^ August 2020. The parameter *b*_*k*_ was always less than 1 during this period indicating a decrease in the reported number of new confirmed cases of the disease during the time segment *k*+1 relative to the time segment *k*. This period was, however, followed by a period of spike in the pandemic up to the time segment beginning 31^st^ August 2020. During this period, the parameter *b*_*k*_ was always more than 1 and even increased to more than 3 during some time segments of this period leading to rapid increase in the reported number of new confirmed cases of the disease. This period of spike in the pandemic was followed by a rapid decrease in the parameter *b*_*k*_ up to the time segment beginning 20^th^ September 2020. The rapid decrease in the parameter *b*_*k*_ during this period resulted in a rapid decrease in the number of new confirmed cases of the disease. However, the pandemic spiked again during the time segment beginning 16^th^ October 2020, but the spike was not large as the parameter *b*_*k*_ was never more than 1.5 during this period.

The next phase of the pandemic appears to have started from the time segment beginning 17^th^ October 2020 and lasted up to the time segment beginning 17^th^ February 2021. This period is characterised by the decrease in the number of daily confirmed cases of the disease as the parameter *b*_*k*_ remained less than 1 most of the time during this period which means that the number of daily confirmed cases of the disease reported during the time segment *k* + 1 was, in general, less than the number of daily confirmed cases of the disease reported during the time segment *k* indicating a digression of the pandemic. However, the parameter *b*_*k*_ increased rapidly during the time segment beginning 18^th^ February 2021 so that, during this period, there had been very rapid increase in the number of daily confirmed cases of the disease reported during the time segment *k* +1 relative to the time segment *k*. This period lasted up to the time segment beginning 16^th^ April 2021 and the parameter *b*_*k*_ reached almost 8 during this period indicating very rapid increase in the number of daily confirmed cases of the disease reported during the time segment *k* +1 relative to the time segment *k*. Although, *b*_*k*_ started decreasing after the time segment beginning in mid-March, 2021 yet, it decreased to less than 1 only after the time segment beginning mid-April 2021 resulting in a decrease in the number of daily confirmed cases of the disease reported during time segment *k* + 1 relative to time segment *k*.

The number of daily reported new confirmed cases of the disease is also influenced by the difference 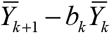. If this difference is greater than 0, then the number of daily reported confirmed cases of the disease is more than that implied by the parameter *b*_*k*_. This, essentially, implies that a proportion of the reported cases are of unknown contact. For example, *b*_*k*_ =1.18857 between time segment beginning 12^th^ June 2020 and immediately succeeding time segment beginning 26^th^ June 2020. This means that the number of confirmed cases of the disease reported on 26^th^ June 2020 (7-days average) should be 1.18857 times the number of confirmed cases of the disease reported on 12^th^ June 2020 (11008) or 13083. However, the number of confirmed cases of the disease, actually reported on 26^th^ June 2020, was 18154. This means that around 5286 cases reported on 26^th^ June 2020 were of unknown contact.

On the other hand, if the difference 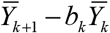 is less than 0, then this implies that all cases of the disease implied by the parameter *b*_*k*_ is not reported. This means that all contacts of the confirmed cases reported during the time segment *k* could not be traced. For example, *b*_*k*_ =2.37482 between the time segment beginning 26^th^ June 2020 and the immediately succeeding time segment beginning 10^th^ July 2020. This means that the number of confirmed cases of the disease reported on 10^th^ July 2020 (7-days average) should be 2.37482 times the number of confirmed cases of the disease reported on 26^th^ June 2020 (18154) or 43111. However, the number of confirmed cases of the disease, actually reported on 10 July 2020, was only 26757. In other words, it appears that around 16372 cases of the disease that might be known contacts of the cases reported on 26^th^ June 2020 could not be reported on 10^th^ July 2020. A part of this gap may be due to reporting delays or inconsistencies, but it also reflects the deficiency in contact tracing and testing of known contacts of already confirmed cases of the disease.

Figure 2 depicts the 7-days of concurrent average of daily reported new confirmed cases of the disease in India beginning from 1^st^ March 2020 through 15^th^ May 2021 along with the fitted values derived from the application of the model (2). It is evident from the figure that the model (2) provides very good fit to the 7-days concurrent average of daily reported number of new confirmed cases of the disease. More specifically, the model effectively captures the very rapid increase in the daily reported number of new confirmed cases of the disease in the recent past which is commonly termed as the second wave of the COVID-19 pandemic in the country.

**Figure 2:**
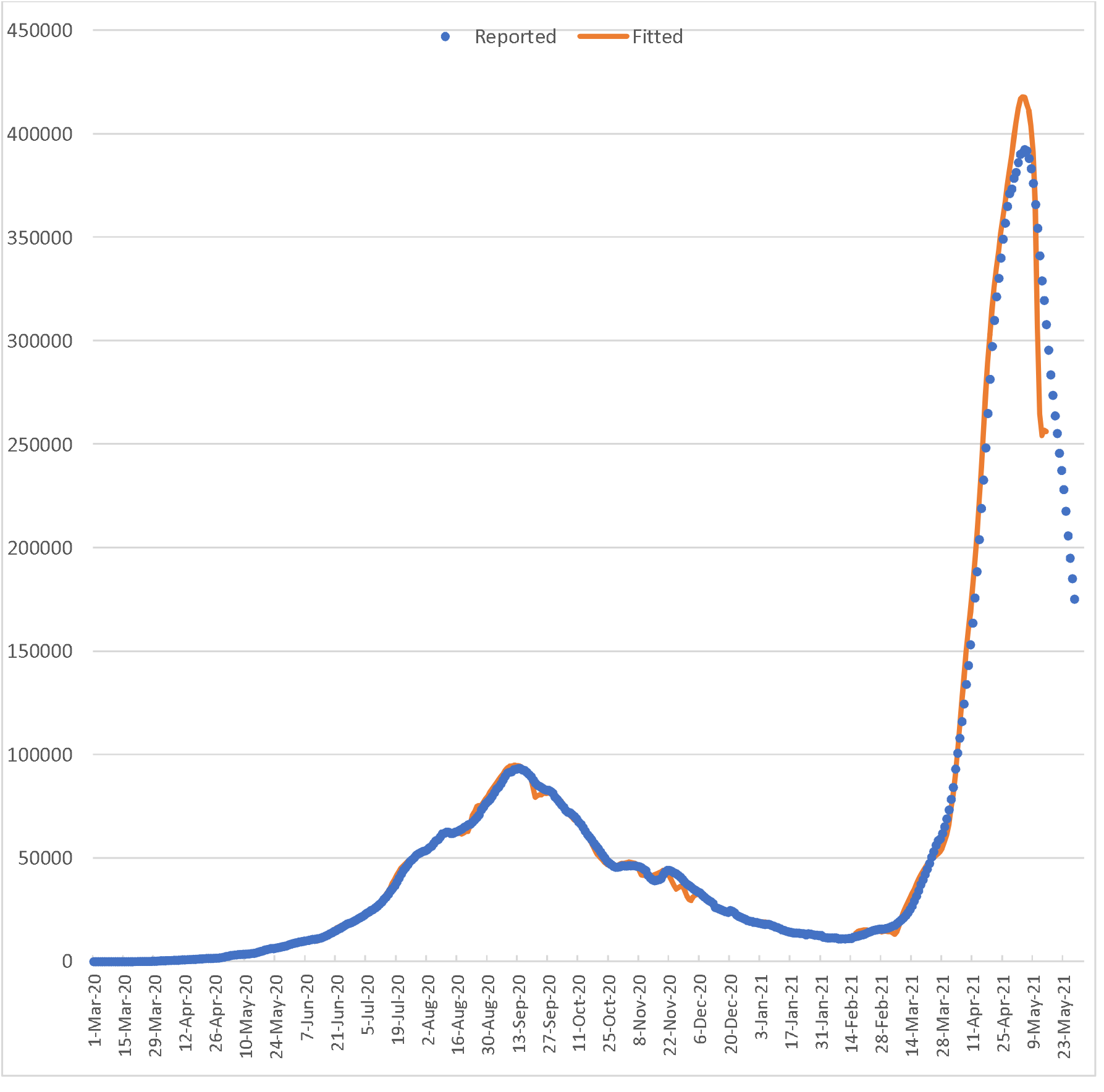
Reported and fitted 7-days concurrent average of reported new confirmed cases of the disease. Source: Authors’ calculations

There has been a very rapid increase in the average daily reported confirmed new cases of the pandemic from 10986 on 8^th^ February 2021 to 396303 on 8^th^ May 2021 (Figure 3). This rapid increase has been the result of both a large base of infected persons and a very high multiplier as reflected through the value of the parameter *b*_*k*_. For example, the parameter *b*_*k*_ is estimated to be 7.28368 between the time segment beginning 22^nd^ February 2021 and time segment beginning 8^th^ March 2021. This means that the number of confirmed cases of the disease reported on 22^nd^ February 2021 (14260) would have multiplied to 103863 on 8^th^ March 2021.

However, the number of confirmed cases of the disease reported on 8^th^ March 2021 was only 19296 which means that 86796 cases of potentially known contacts of the cases reported on 22^nd^ February 2021 could not be reported on 8^th^ March 2021. This unreported pool of confirmed cases of the disease appears to have contributed to increasing the daily reported confirmed cases of the disease even after the multiplying factor *b*_*k*_ decreased to less than 1. For example, the parameter *b*_*k*_ between the time segment beginning 7^th^ April 2021 and time segment beginning 21^st^ April 2021 is estimated to be 0.44367 which means that the reported number of cases on 7^th^ April 2021 (124476) would have decreased to 55226 on 21^st^ April 2021. However, the number of confirmed cases reported on 21^st^ April 2021 was 309858. This means that 270521 cases of unknown contact were reported on 21^st^ April 2021. Had there been effective tracing of contacts of all confirmed cases of the disease reported in the past, the reported umber of confirmed cases of unknown contact would not have been so large.

The parameter *b*_*k*_ is the key factor in deciding the pace of the progression of the pandemic. During the time segment beginning 8^th^ February 2021 through the time segment beginning 17^th^ February 2021, *b*_*k*_ was negative indicating the confirmed cases of the disease reported during the time segment beginning 17^th^ February 2021 were negatively correlated with the confirmed cases of the disease reported during the time segment beginning 3^rd^ February 2021. It, however, increased to more than 1 during the time segment beginning 19^th^ February 2021 indicating that the second wave of the pandemic started on 19^th^ February 2021. There has been a very rapid increase in *b*_*k*_ during the time segment beginning 1^st^ March 2021 through the time segment beginning 8^th^ March 2021 when *b*_*k*_ increased to 7.28 which implied that 100 infected persons during the time segment beginning 22^nd^ February 2021 infected, on average 728 persons during the time segment beginning 8^th^ March 2021 leading to a very rapid increase in the number of daily reported confirmed cases of the disease. The value of *b*_*k*_ started decreasing after the time segment beginning 8^th^ March 2021 and decreased to 1.74 during the time segment beginning 20^th^ March 2021 but increased again to almost 3.6 during the time segment beginning 2^nd^ April 2021, after which, it started decreasing. It is obvious that very rapid increase in the average daily reported number of new cases of the disease during the second wave of the pandemic may be attributed to very rapid increase in the parameter *b*_*k*_ during the time segment beginning 1^st^ March 2021 through the time segment beginning 10^th^ April 2021. Moreover, a large number of known contacts of cases reported during the immediately preceding time segments could not be reported so that the number of cases of unknown contact increased substantially. It may, however, be noted that despite the parameter *b*_*k*_ turned less than 1 after 17^th^ April 2021, the reported number of confirmed cases of the disease continued to increase till 8^th^ May 2021 because of the large base of the already infected persons. It was only after time segment beginning 8^th^ May 2021 that the reported number of confirmed cases of the disease started decreasing.

Figure 2 empirically validates the approach adopted in this paper in analysing and describing the progression of the pandemic. It may also be noted that 7-days concurrent average of daily reported number of new confirmed cases of the disease may be forecasted in the immediate future by projecting the parameter *b*_*k*_. It may, however, be noted that the trend in parameter *b*_*k*_ is highly volatile so that long-term projection of the parameter *b*_*k*_ does not appear to be a feasible proposition. The very high volatility of the parameter *b*_*k*_ probably and so obviously explains the reason why most of the existing approaches have not been successful in forecasting the progression of the pandemic.

## Conclusions

This paper has followed a data driven approach to analyse and describe the COVID-19 pandemic in India. The approach does not make any assumption about the progression of the pandemic. It is based on the relationship between the number of confirmed cases of the disease reported during a time segment of 14-days with the number of confirmed cases of the disease reported during the immediately preceding time segment of 14-days as it is well known that a person tested positive for Novel Coronavirus remains infective for a period of 14-days only. Beyond the period of 14-days, the infected person contributes little to the progression of the pandemic.

The present analysis suggests that two factors may be attributed to the progression of the pandemic in the country. The first, obviously, is the multiplicity of the reported confirmed cases of the disease which is measured in terms of the parameter *b*_*k*_ of the model adopted in the present analysis. The higher this multiplicity the higher the momentum for the progression of the pandemic. The present analysis shows that the multiplicity of the reported confirmed cases of the disease was very high in the beginning of both first and second waves of the pandemic. Moreover, the duration when the multiplicity of reported confirmed cases of the disease was exceptionally high was longer at the time of the second wave compared to the first wave. This exceptionally high multiplicity resulted in a large pool of infected persons that contributed to a rapid increase in the reported number of confirmed cases of the disease even at low levels of multiplicity. Reasons for the increase in multiplicity may be traced in the new strains of virus that attacked the country.

The second factor in the progression of the pandemic in the country appears to be the inability to trace and isolate all contacts of daily reported number of confirmed cases of the disease which also resulted in a large number of reported confirmed cases of the disease with unknown contacts. This has particularly been the case at the time of the second wave of the pandemic leading to very rapid increase in the reported number of confirmed cases. Our analysis suggests that the reported number of confirmed cases of the disease with unknown contacts still remains very high and this very large pool of infected persons without known contact has primarily been responsible for exceptionally high reported number of new confirmed cases of the disease. The progression of the pandemic would have been radically different if all contacts of reported number of new confirmed cases of the disease would have been traced and infected contacts would have been isolated when the multiplicity of the reported number of new confirmed cases of the diseases would have started increasing because of the attack of the new strain of the Novel Corona virus. The progression of the pandemic would have been radically different if all contacts of reported number of confirmed cases of the disease would have been traced and infected contacts would have been isolated when the reported number of new confirmed cases of the diseases had started multiplying at a rapid rate because of the attack of the new strain of the Novel Coronavirus.

Our analysis also suggests that forecasting the progression of the pandemic on the basis of the reported number of new cases of the disease is possible by projecting the trend in the parameter *b*_*k*_. It may, however, be noticed that the trend in *b*_*k*_ is highly volatile so that projection for a short duration only is meaningful. However, from policy and programme perspective, the analysis suggests monitoring the trend in *b*_*k*_ closely and initiate appropriate control measures as soon as the parameter *b*_*k*_ crosses the limiting value of 1. In other words, an early warning system based on *b*_*k*_ may be established to prevent the looming third wave of the pandemic in the country.

## Data Availability

All data used in the analysis are available online:
www.covid19india.org

## Contributions

ARC conceptualised the study, developed the model and carried out calculations.

BPS carried out the literature and model review and read the manuscript.

RS read and edited the manuscript.

## Funding

There is no funding.

## Competing Interests

There are no competing interests.

---

**Table 1.**
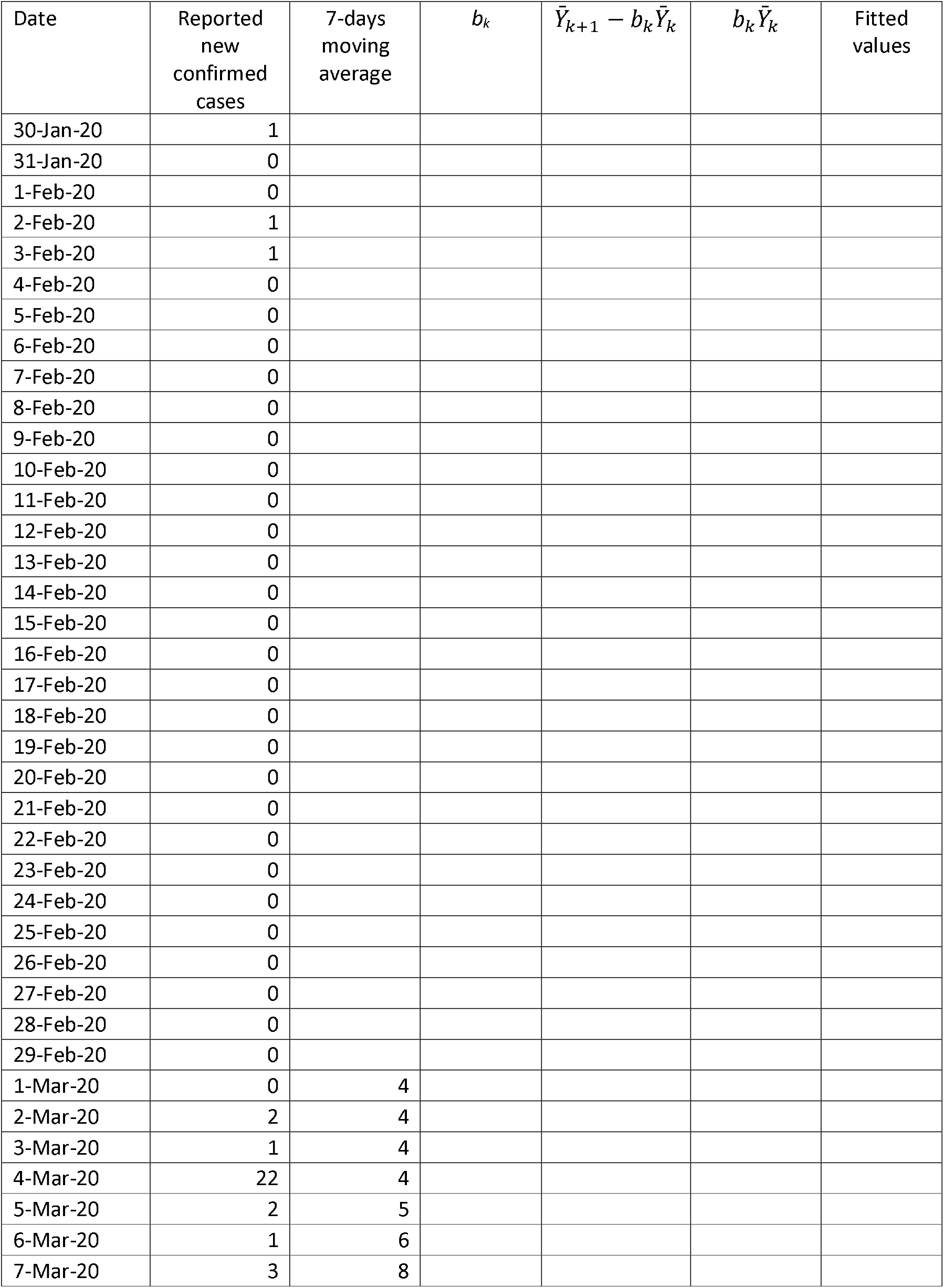
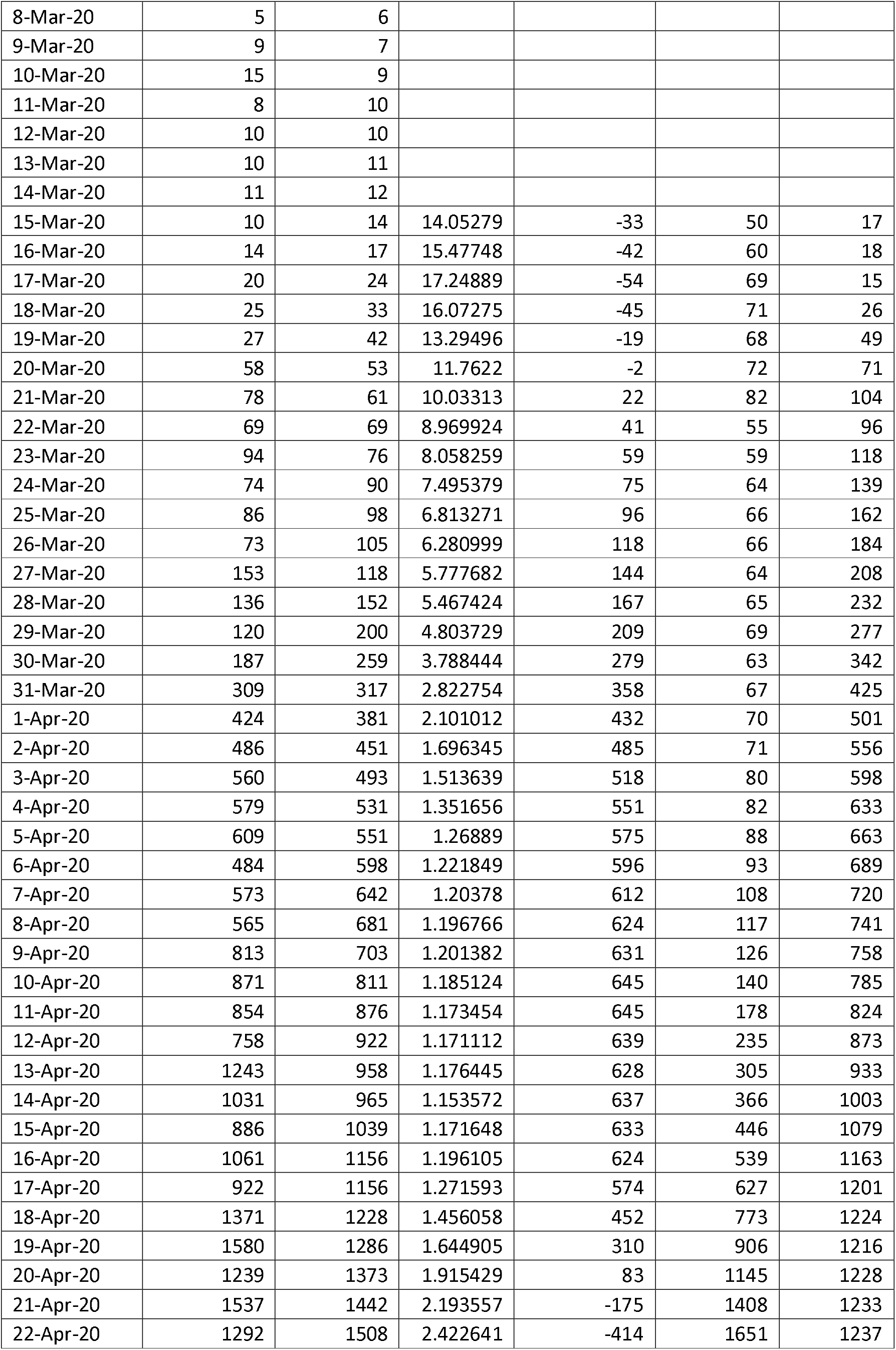
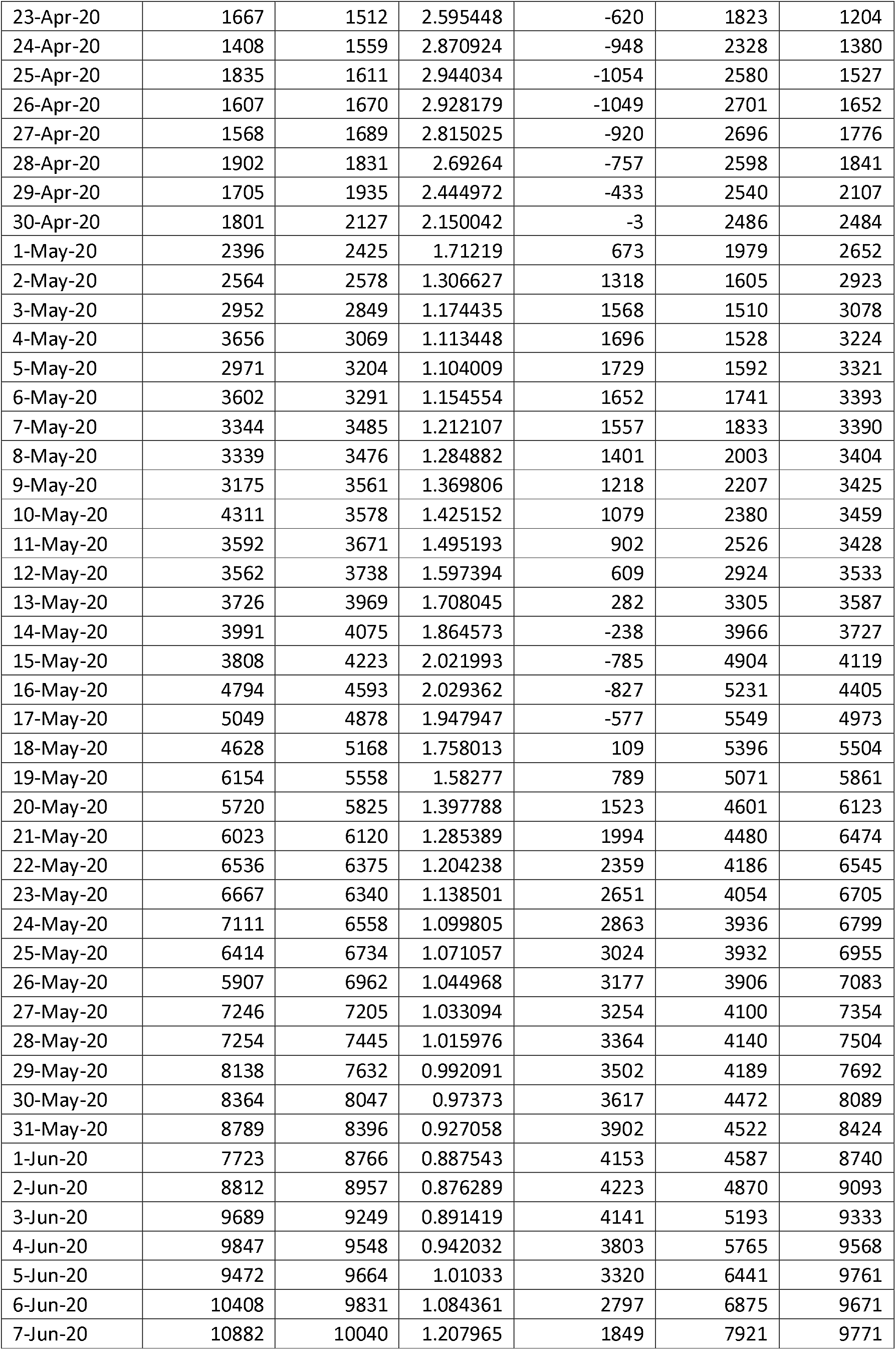
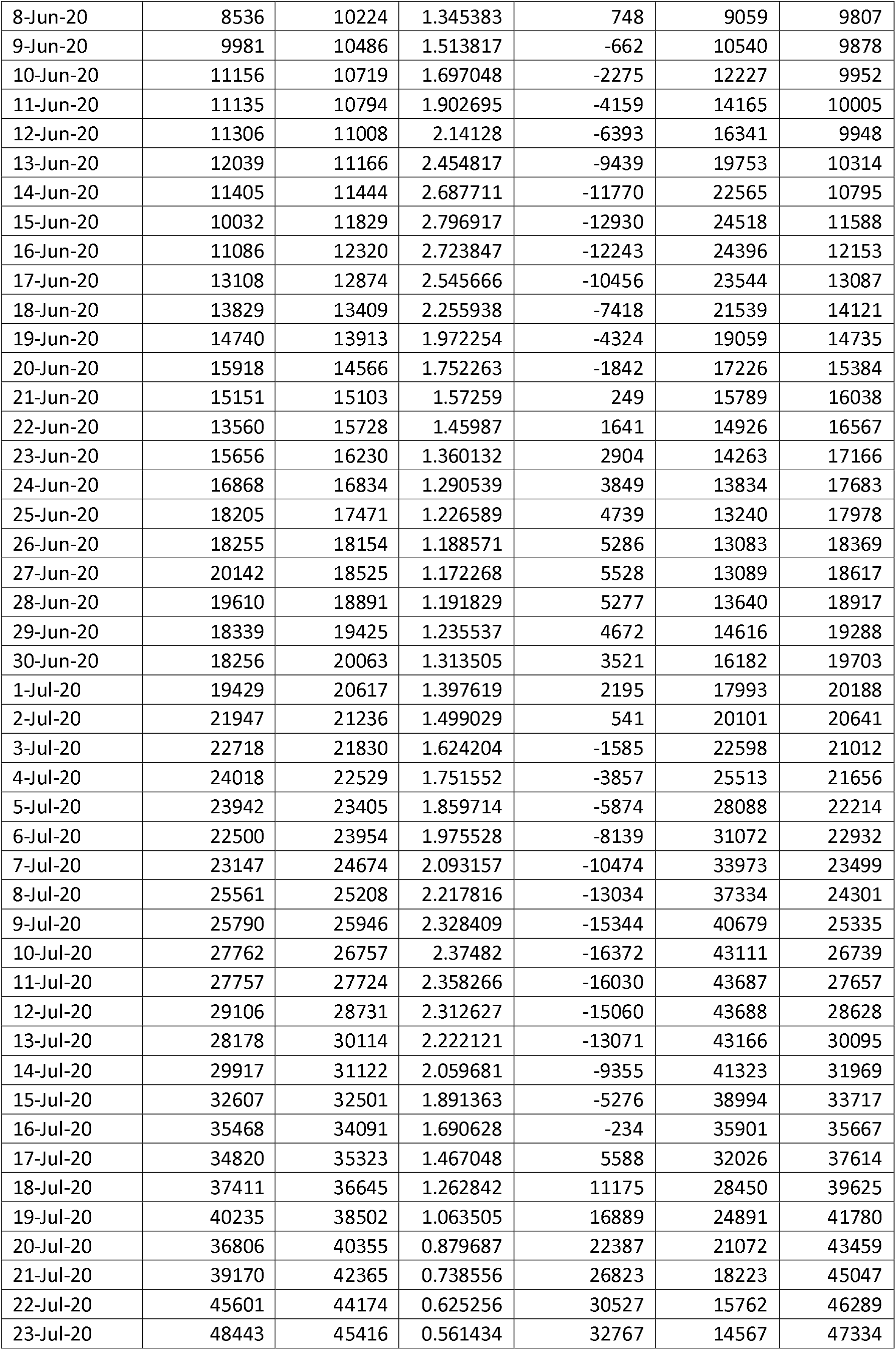
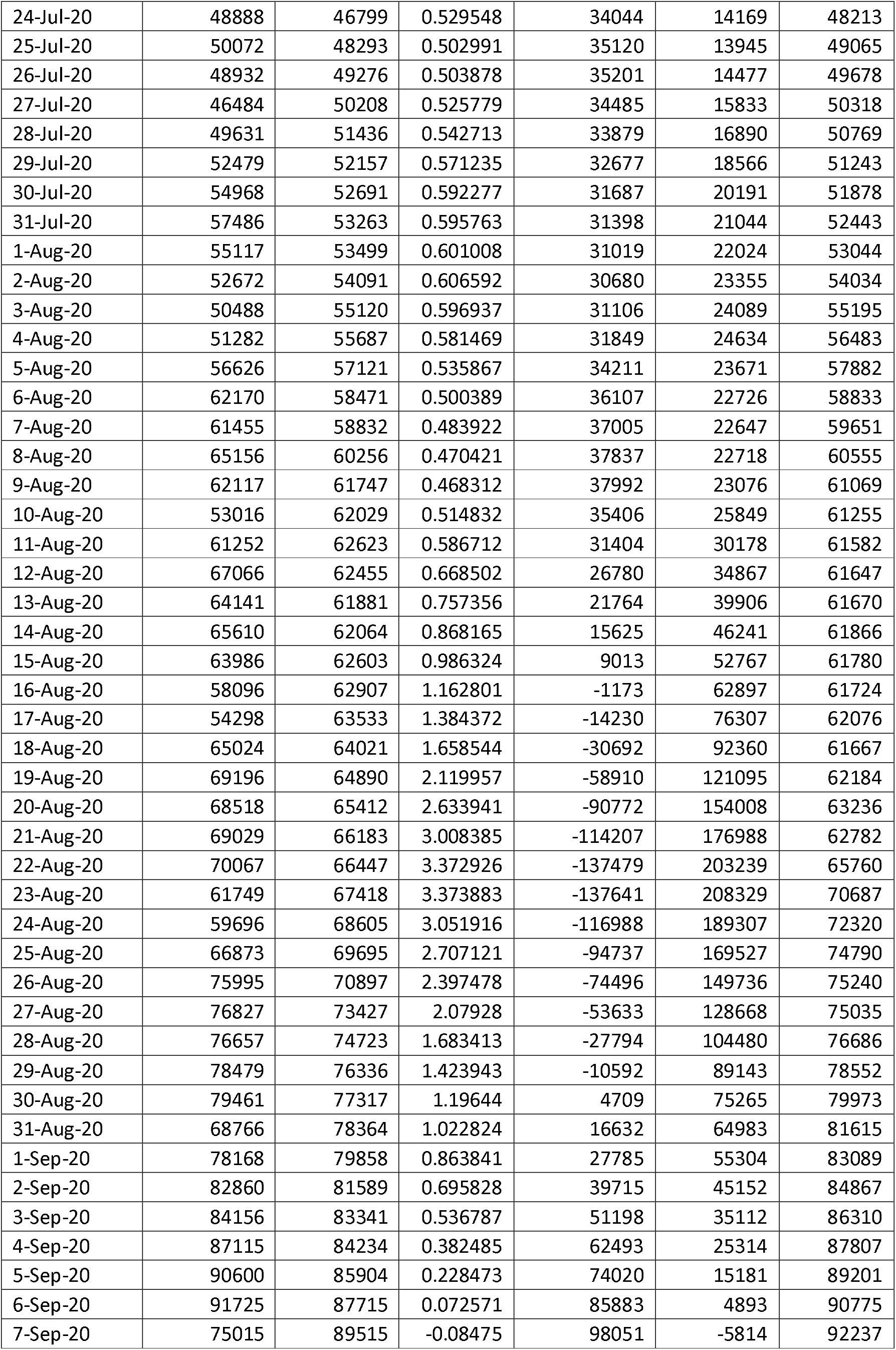
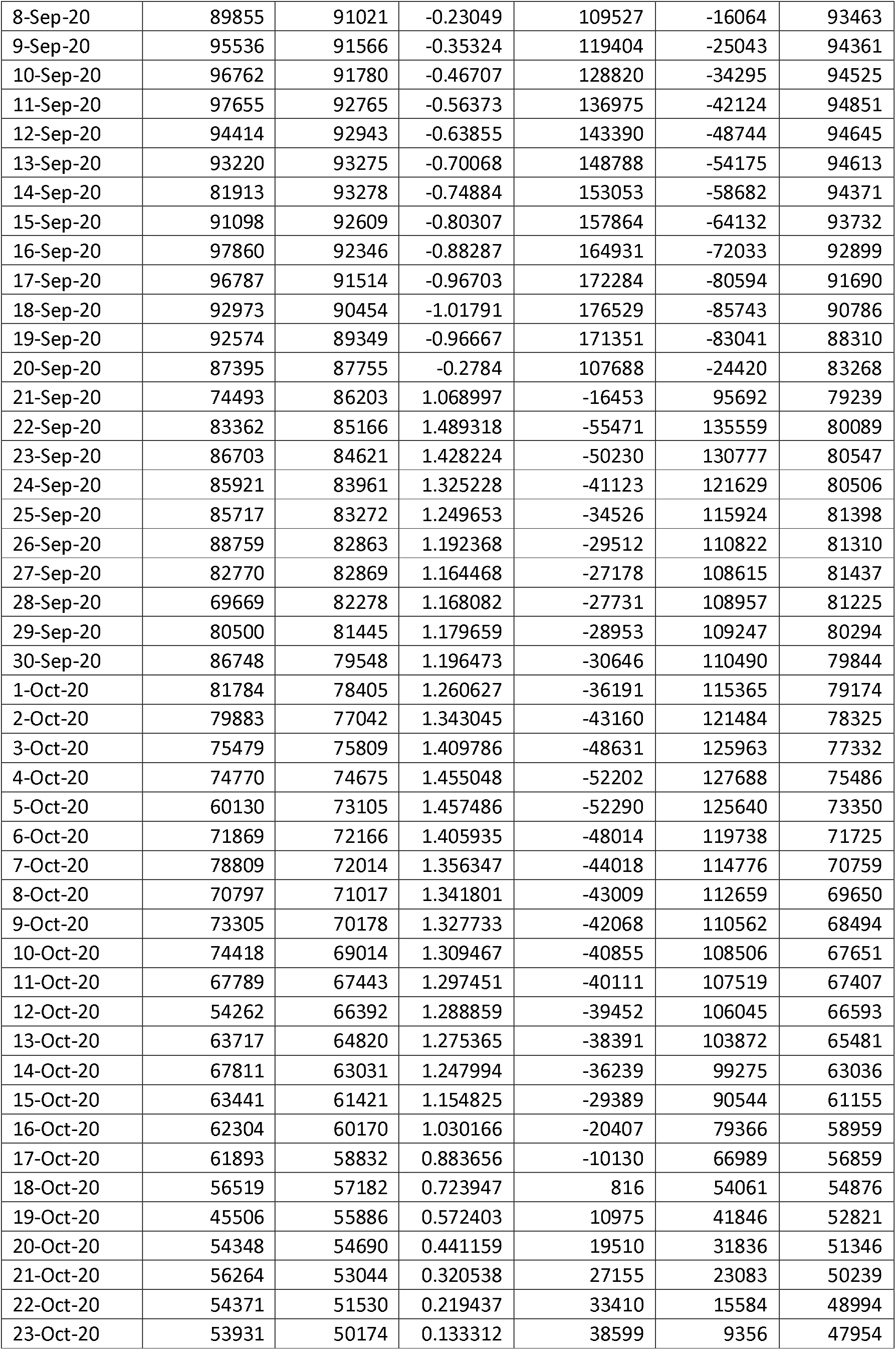
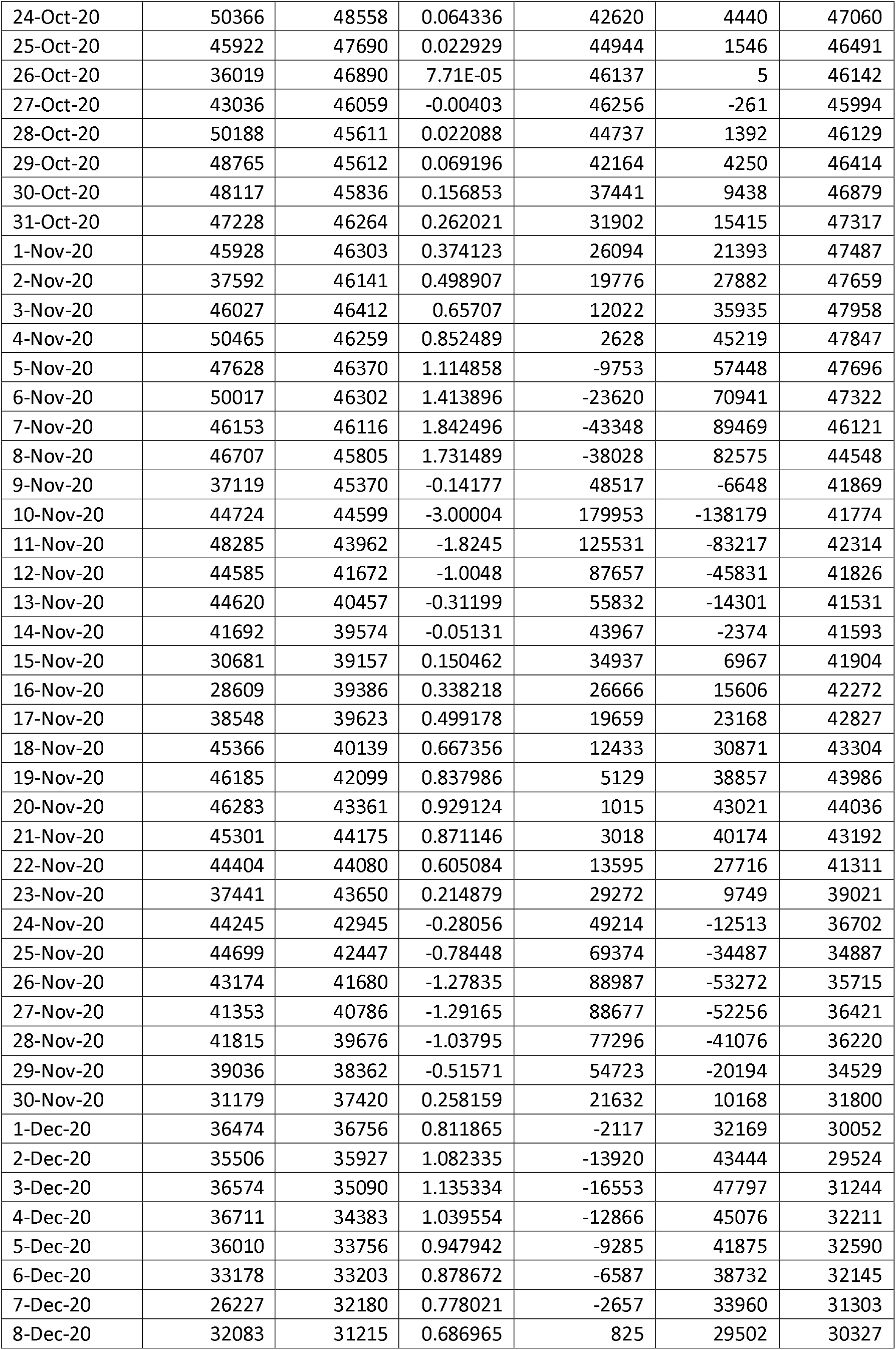
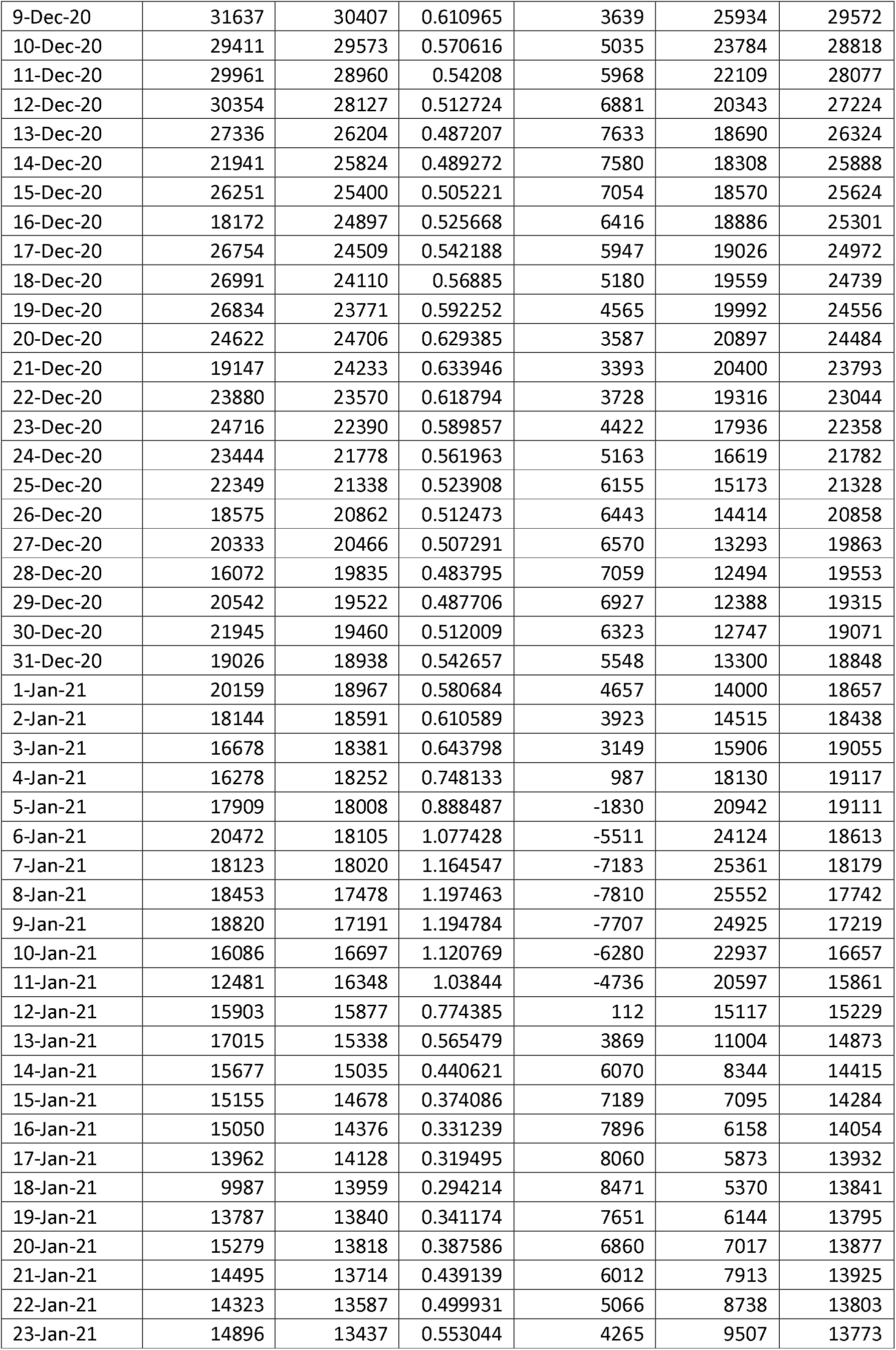
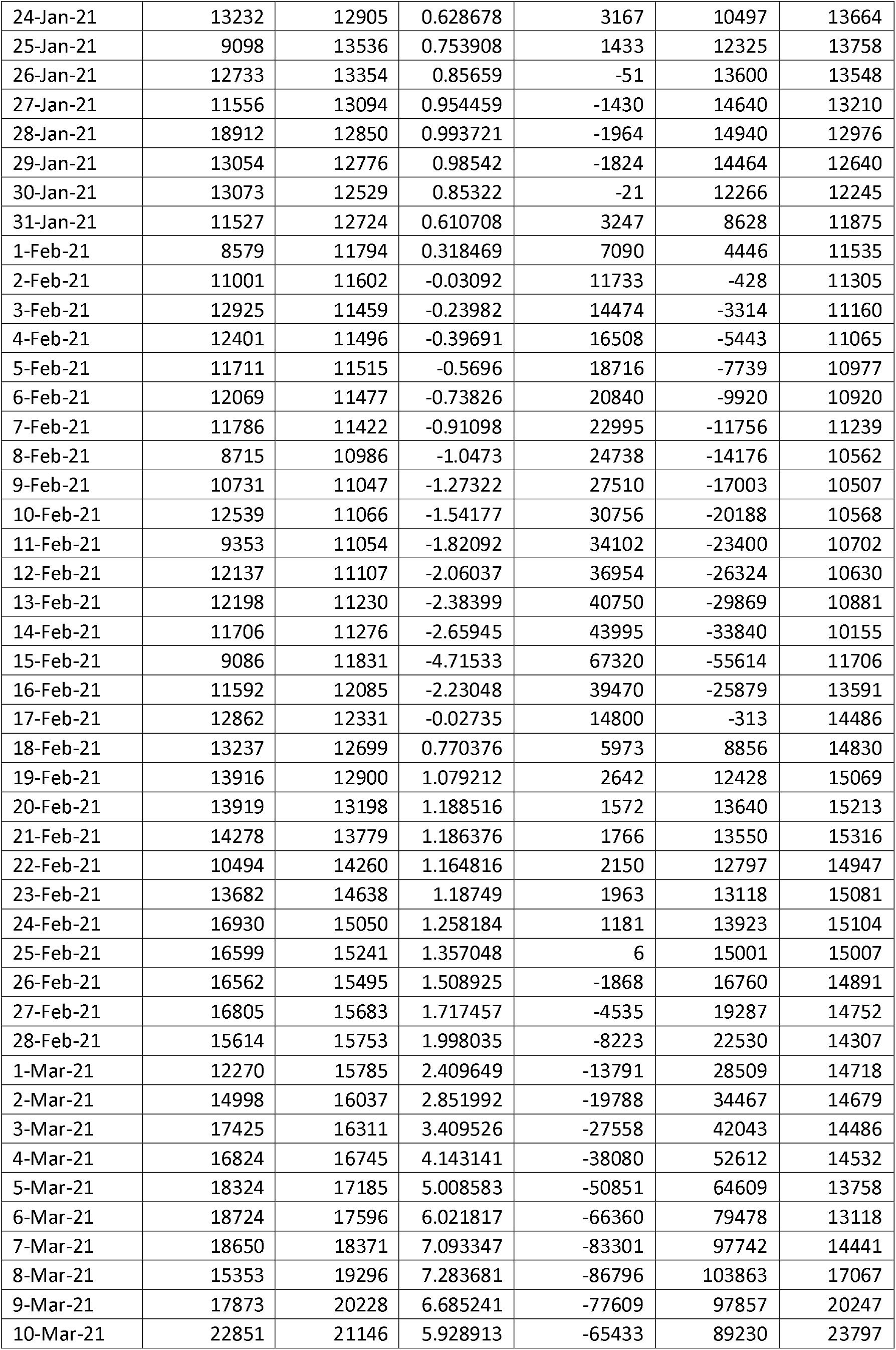
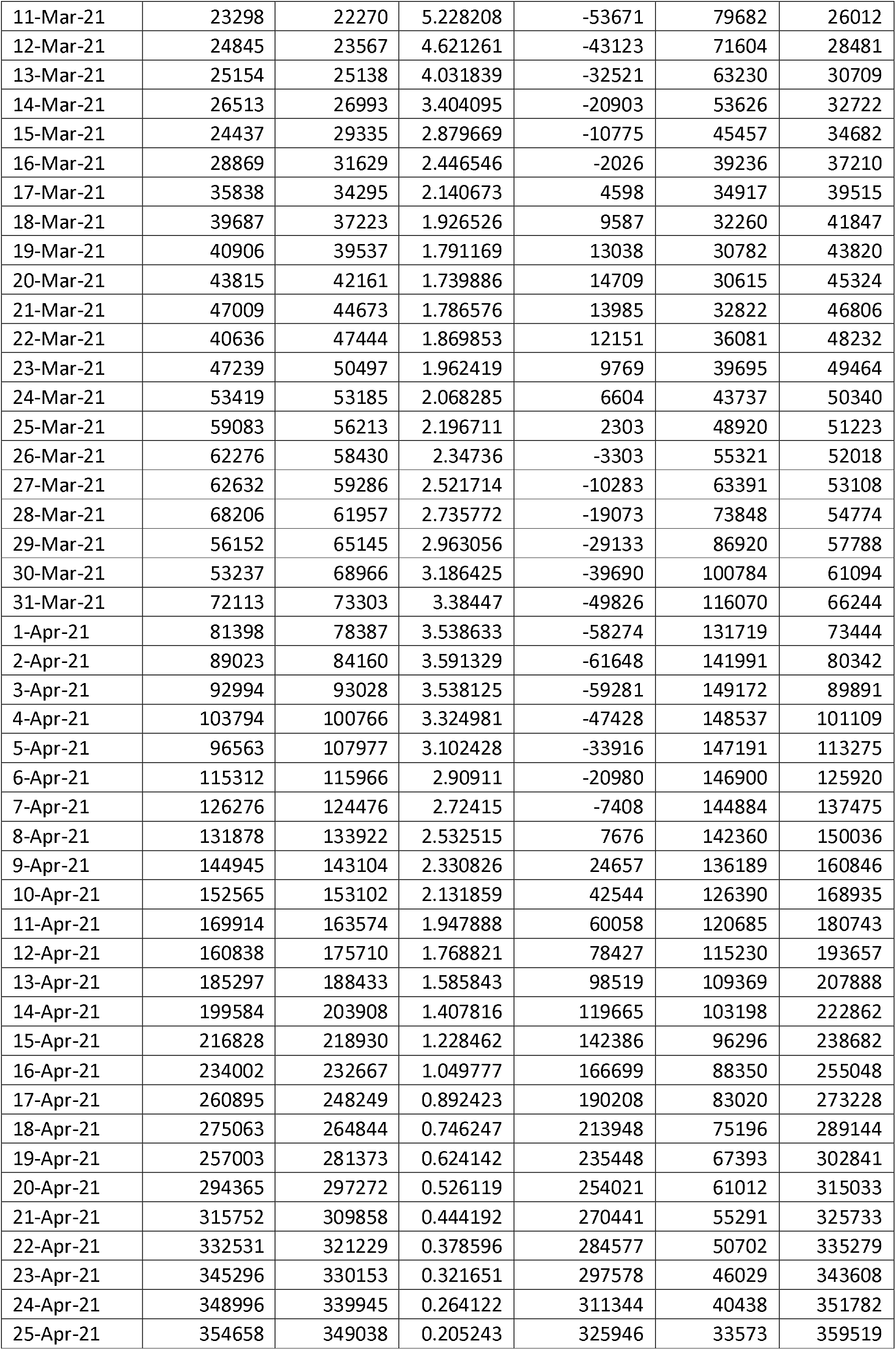
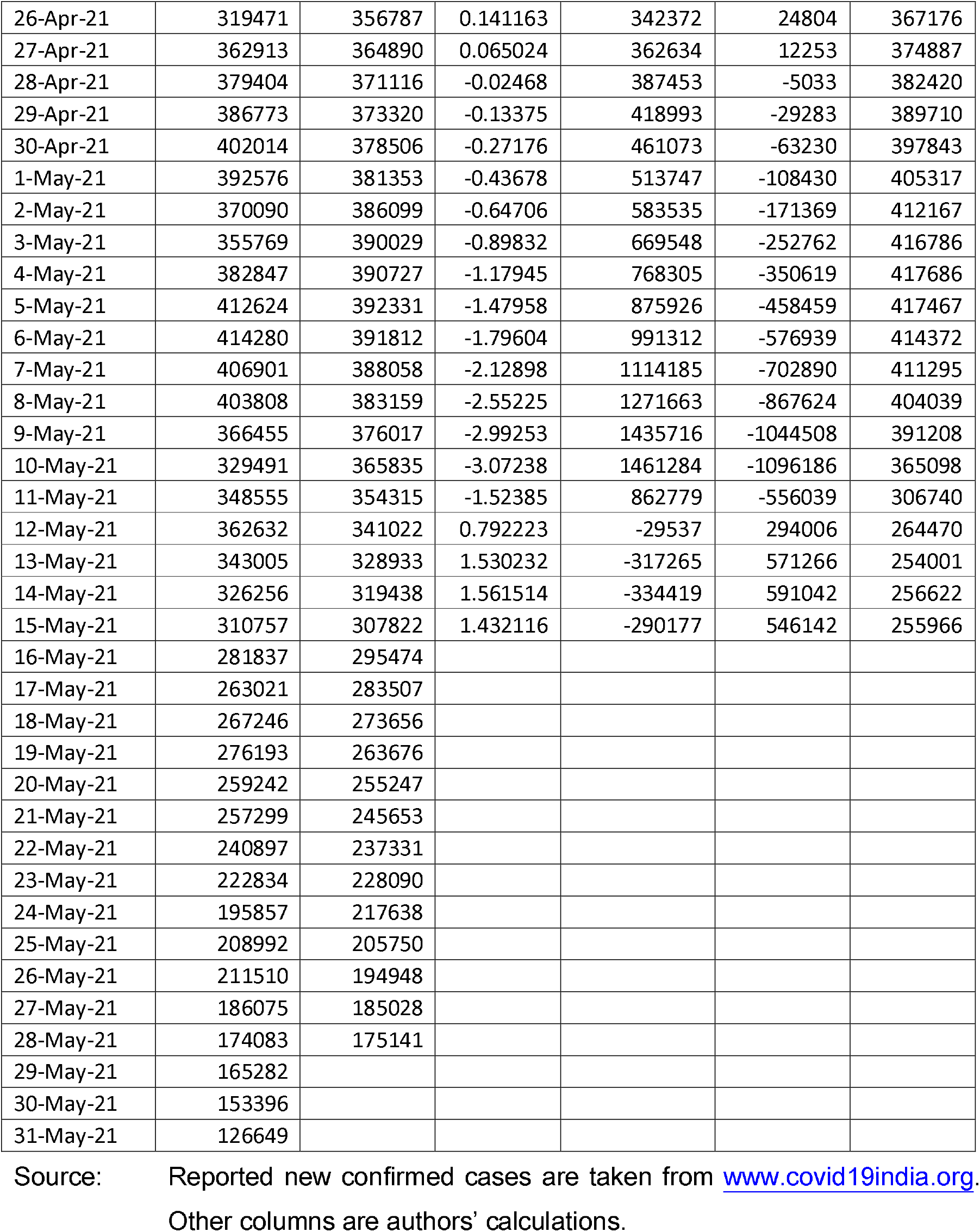
Observed and fitted values of 7-days average of new confirmed cases of COVID 19 in India.

## Notes

### Competing Interest Statement

The authors have declared no competing interest.

### Author Declarations

Institutional oversight body of MLC Foundation.

